# Geographical targeting of active case finding for tuberculosis in Pakistan using artificial intelligence software: a qualitative study embedded within the SPOT TB trial

**DOI:** 10.64898/2026.08.18.26360657

**Authors:** Alina Shahid, Abdullah Latif, Alizeh Faran, Amna Mahfooz, Syed Mohammad Asad Zaidi, Wasim Ahmed, Nainan Nawaz, Tahira Ezra Reza, Faran Emmanuel

**Author notes:** Corresponding author: Alina Shahid.

## Abstract

**Background:** Tuberculosis (TB) remains a critical public health challenge in Pakistan. The SPOT-TB trial evaluated MATCH-AI; an AI tool designed to geographically target active case finding (ACF) by identifying sites for screening TB. Qualitative study was conducted to examine field team and stakeholder experiences to understand the human, organizational, and contextual factors effecting implementation.

**Methods:** Five sub-recipients (SRs) were randomly selected; two districts per SR based on certain selection criteria. Thematic analysis was conducted on thirty In-Depth Interviews (IDIs) and two Focus Group Discussions (FGDs), guided by the Socio-Technical Systems (STS) framework.

**Findings:** Themes included (1) MATCH-AI as a useful tool, (2) operational and contextual challenges, (3) challenges of the staff, (4) organizational readiness, and (5) stakeholder engagement across hierarchy. The staff valued MATCH-AI for reducing bias and external pressure, and it identified TB cases in previously overlooked areas. Local knowledge of staff was crucial as the AI didnot account for operational barriers and contextual issues in certain areas. Weak infrastructure, and inconsistent stakeholder engagement, the system lacked the readiness needed for a new technology to make optimal impact. Understanding of how MATCH-AI functioned varied across hierarchical levels diminishing the sense of ownership among field staff.

**Interpretation:** MATCH-AI holds genuine potential to systematize TB screening and reduce selection bias. Yet it cannot replace the contextual intelligence of field staff like knowledge of community trust, gender norms, and security realities. Effective implementation demands reliable infrastructure, meaningful stakeholder engagement, and field staff orientation. AI integration succeeds only when technical solutions align with human and organizational readiness.

## Introduction

Despite decades of progress, tuberculosis (TB) remains a leading cause of morbidity and mortality globally. Pakistan has the world’s 5th highest TB burden, and over a third of all incident cases annually are undiagnosed or not reported.^1^ To increase case-detection for TB, Pakistan implemented the use active case finding (ACF) through mobile X-ray–based screening, rolling it out as a programmatic intervention in 2017.^2^ ACF screening events are known as “camps” in Pakistan and involve activities like community mobilization, participant recruitment, screening, and sputum collection for diagnostic testing. Camp sites are selected based on factors including TB notification data, partnerships with community leaders and local organizations, and field staff experience. ^2^ However, yields from ACF are lower than expected, prompting the search for strategies to improved case-detection.

Previous studies in Pakistan have shown that TB risk varies across regions, suggesting that screening is more effective when focused on high-risk areas. The 2019 TB Joint Programme Review Mission (JPRM), led by the Government of Pakistan with WHO and partners, also reported lower-than-expected ACF yields and recommended using geographic targeting in areas with high TB burden.^3^ In response, the National TB Control Program (NTP) introduced MATCH- AI (Mapping and Analysis for Tailored Disease Control and Health System Strengthening– Artificial Intelligence).^4^ This decision-support tool combines routine TB program data, ACF data, and local contextual information to identify areas with a high risk of TB using Bayesian modeling. The original MATCH framework, developed by the KIT Royal Tropical Institute in 2017, has been used in several high-burden countries to identify areas with high estimated TB burden but low case detection. It was later adapted into an AI-based tool by EPCON and piloted by the NTP and Mercy Corps in selected districts of Pakistan in 2019.^4^

Hence, a pragmatic stepped-wedge trial (SPOT-TB) was conducted between August 2023 and September 2024 to evaluate whether AI-guided geographic targeting improved the detection of bacteriologically confirmed TB compared with conventional site selection methods. Across 3,936 screening camps and over 269,000 individuals screened, the AI strategy did not significantly improve TB detection in the intention-to-treat analysis. Compliance with the exact locations generated by MATCH-AI was, however, somewhat low: of the camps in the intervention arm, only around half could be verified as having actually been conducted at or near the AI-recommended site. When restricted to this subset of camps with confirmed high compliance, AI-guided targeting achieved a 32% higher yield of bacteriologically confirmed TB cases compared with routine site selection, suggesting that the lack of an overall effect in the intention-to-treat analysis may be driven, at least in part, by incomplete compliance to the tool’s recommendations during implementation.^5^

While these innovations highlight the potential of AI to strengthen targeting in TB control programs, the on-ground implementation of AI based technology still remains complex. Field implementation of AI supported programs especially in the low- and middle-income countries (LMICs) is often challenged by structural and systemic barriers including infrastructure, workflow, trust and governance.^6–9^ Hence, complementing the trial, we conducted a qualitative study to see how MATCH-AI was used in practice, including field staff adherence to AI recommendations, trust and acceptability of the tool, and operational factors influencing its implementation.

## Methodology

### Study Design and Setting

We conducted a qualitative study during the final quarter of the rollout of the SPOT TB trial (June–September 2024). This study reports key aspects of the research process according to the Consolidated Criteria for Reporting Qualitative Research (COREQ) guidelines.^10^ As shown in fig 1, MC is a principal recipient and implements camps in collaboration with implementing partners, also known as Sub-Recipients (SRs). The study was conducted in 14 districts in four provinces of Pakistan.

**Figure 1.**
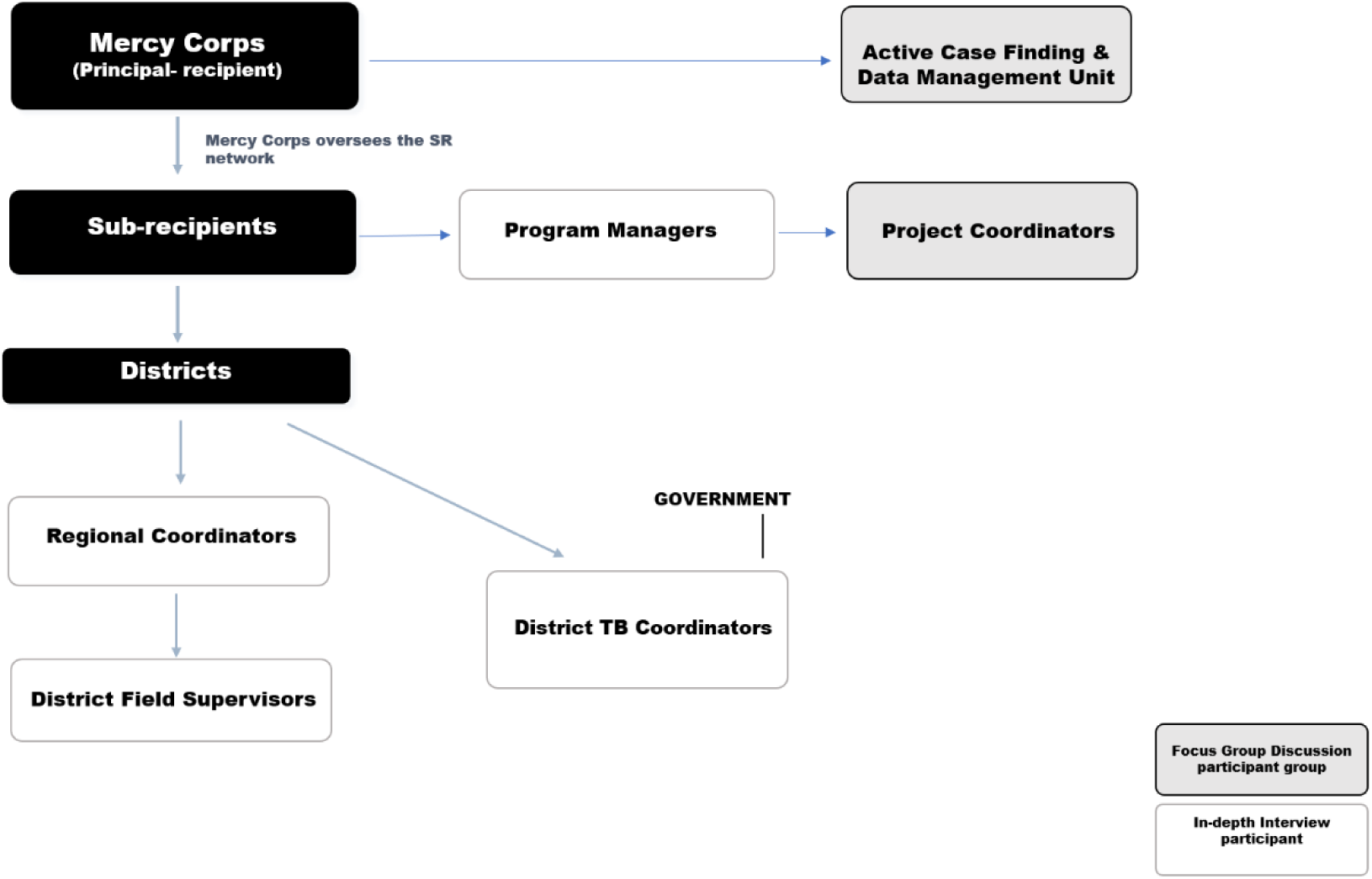
Organizational hierarchy and study participants

**Figure 2.**
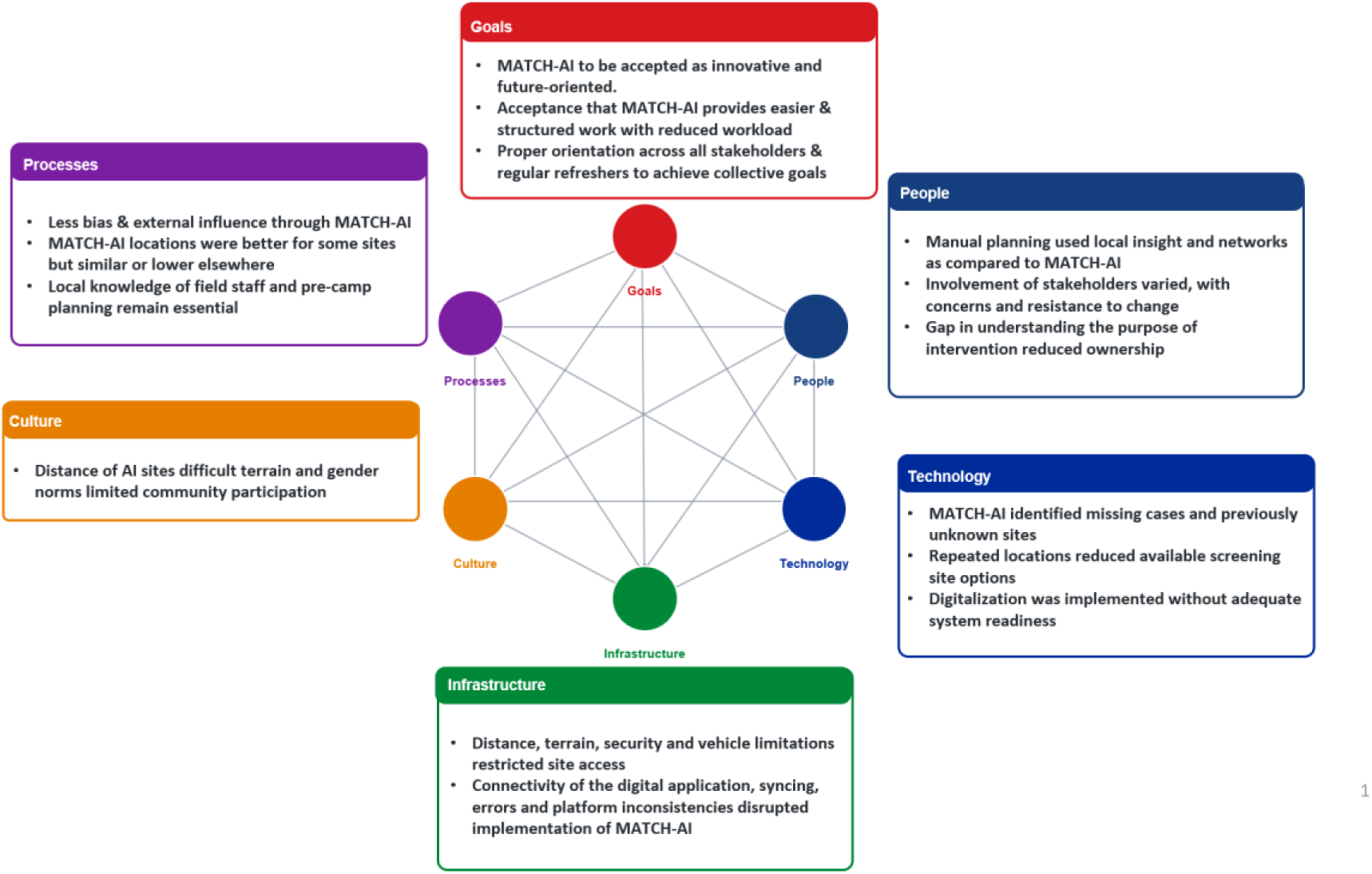
MATCH-AI Implementation Through a Socio-Technical Systems Lens

### Participants

There were a total of nine SRs working with MC in mutually exclusive districts. Five SRs were randomly selected, among whom staff from two districts each were enrolled into the study: one from a district with high compliance to MATCH-AI recommendations, and one from a district with low compliance (table 1). Data from the previous four months were reviewed to identify districts that had completed at least three months in the intervention arm.

**Table 1.**
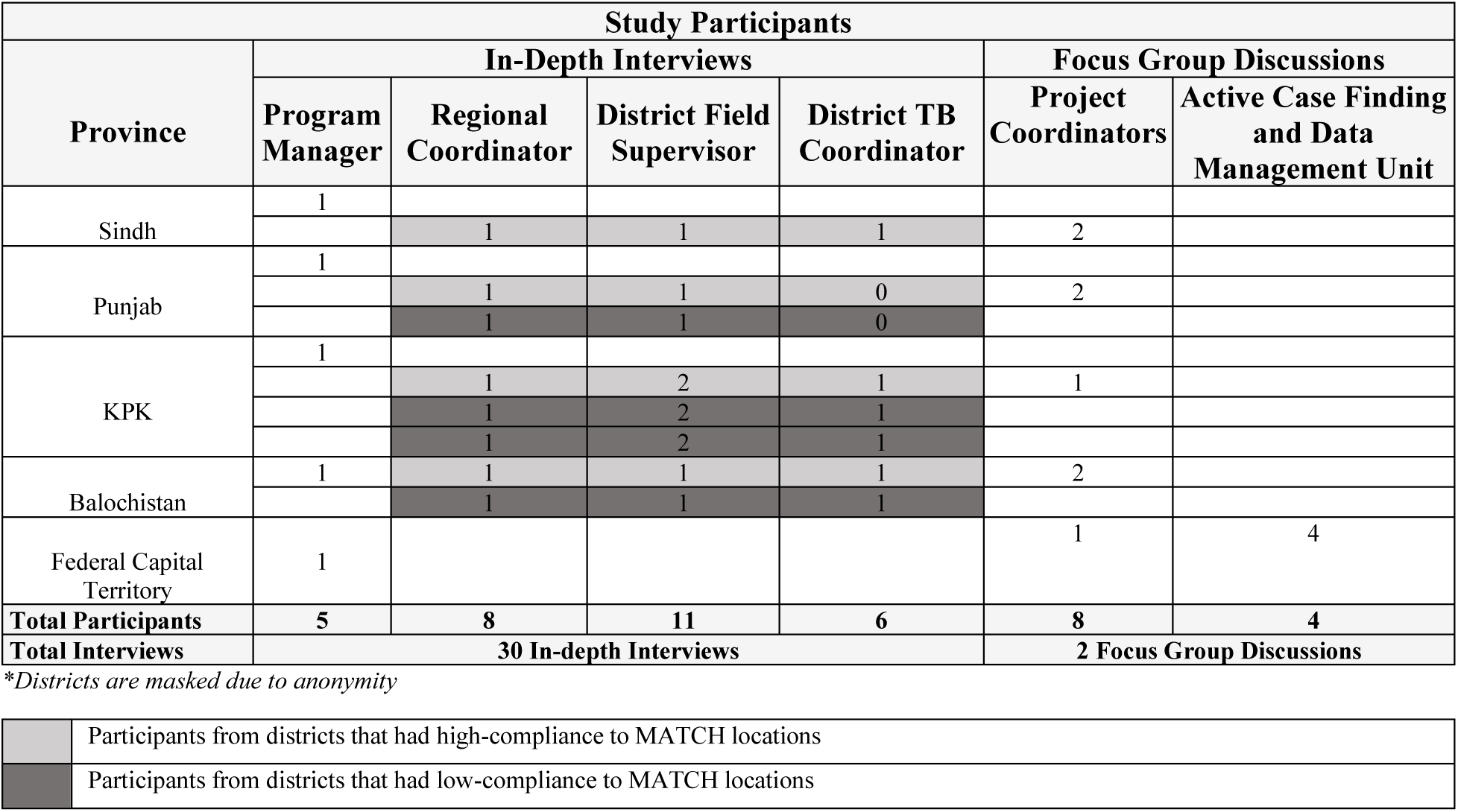
Overview of study participants

From the list provided by MC of all the project staff, individuals with greater implementation experience were identified. The Project Managers (PM) who lead the ACF implementation from the respective SR were interviewed. Then, in each district, interviews were conducted with one Regional Coordinator (RC), one or two District Field Supervisors (DFSs), one District TB Coordinator (DTC) who is a government official. Two DTCs declined participation, resulting in a total of 30 in-depth interviews (IDIs). Data collection continued until saturation was reached, at which point two districts from two different provinces were excluded from further interviews.

One FGD was held with four staff members from the ACF Unit and DMU (Data Management Unit) team at the MC Head Office, as they were directly involved in managing ACF implementation, generating and disseminating MATCH-AI site selections, and receiving field-level feedback and complaints. The second FGD included eight Project Coordinators representing all SRs, who oversee district-level teams and were responsible for coordinating implementation across sites under their respective SRs. (fig 1) The consent was obtained and the study was briefed prior to commencement of interviews. Anonymity of participants was maintained through the use of identification codes.

### Data Collection

Four topic guides were developed in line with study objectives. The guides were initially piloted with three participants and appropriately modified. While broadly similar, the separate guides were tailored to the participants’ scope of work: (i) IDIs with PMs/RCs/DFSs, (ii) IDI with DTCs, and (iii) FGD with data managers, (iv) FGD with PCs, covering topics of camp planning, location and yield, digitalization, and data use.

Two female researchers with expertise in public health (AS and AM), a Qualitative Research Officer and Project Manager conducted all in-person interviews across districts. The interviewers had prior knowledge of the challenges faced by the field staff through verbal feedback from the main trial.^5^ This familiarity informed the development of the interview guide and helped inform probing during interviews to better complement the findings of the trial. IDIs lasted 30–60 minutes and FGDs 70–90 minutes. In total, 30 IDIs and 2 FGDs were conducted (13 by AS, 13 by AM, and 6 jointly). In all instances, at least one of the trial investigators accompanied the researchers, providing support with note taking and observation. There were no incomplete interviews. Sessions were conducted in Urdu, audio-recorded with informed consent, anonymized, and stored securely.

### Theoretical Framework

To help inform the interview guides and the data analysis, we used the Socio-Technical Systems (STS) framework. This theoretical model conceptualizes technology implementation as the result of interactions between technical, social, organizational, and environmental components rather than as a purely technical process.^11,12^ Unlike technology adoption frameworks, STS explicitly recognizes the interdependence of people, technology, culture, processes, goals, and infrastructure.^11^ This makes it particularly well suited for understanding the implementation of MATCH-AI within the complex operational environment of community-based TB active case finding.

### Data Analysis

The study was informed by an interpretivist, contextualist methodological orientation, recognizing that participants’ experiences and accounts are shaped by their organizational, social, and implementation contexts. Data were analyzed using reflexive thematic analysis following the six-phase approach described by Braun and Clarke, which involves familiarization with the data, generation of initial codes, searching for themes, reviewing themes, defining and naming themes, and producing the final report.^13^ Analysis was conducted using a hybrid inductive-deductive approach, whereby coding was informed by the study objectives and sensitized by the STS framework, while allowing for patterns and meanings not anticipated a priori to be developed from the data. Coding was conducted in ATLAS.ti by two researchers, who engaged iteratively with the transcripts and discussed interpretations throughout the analytic process. Initial codes were organized into categories and subsequently interpreted through six interconnected components adapted from the STS framework. These components provided a coherent way to situate participants’ experiences within the sociotechnical context of integrating MATCH-AI into the ACF programme (table 2).^11,12^ Details on how the STS constructs were interpreted for this study are provided in supplementary file 5. Themes were developed through iterative engagement with the coded data, reviewed in relation to the dataset and study objectives, and refined through discussions among the research team. Five overarching themes were developed. Participant quotations are presented verbatim unless minor rephrasing was undertaken to improve clarity while preserving the original meaning. Triangulation across roles (DFS, DTC, RC, PM, PC, data teams from ACF and DMU), data sources (IDIs, FGDs, field notes), and provinces enhanced credibility. Participants were invited to review a summary of the study findings and provide comments regarding their accuracy and interpretation. Overall, participants indicated that the summary was broadly accurate and reflected their experiences well.

**Table 2.** Results

| Codes | Categories | Themes | Categories mapped against STS Framework Constructs |
| --- | --- | --- | --- |
| Welcomed MATCH, Positive reaction to MATCH-AI, appreciating software, first to implement MATCH-AI globally, future is technology | Positive Reception | **MATCH-AI as a useful tool** | Goals |
| MATCH makes work easier, Workload decreases with MATCH, work became structured, Data-driven camp selection | Efficiency and Ease |  | Goals |
| Public registers for TB locations, Data-backed site selection yields results, Find missing cases with MATCH, AI gives multiple choices, Systemic approach of AI, Site we never knew existed gave cases | Data-Driven Decisions |  | Technology |
| No favoritism in AI, Lack of external influence in MATCH, bias is minimized | Reduced Bias |  | Process |
| Better yield with AI, Good yield from new sites, Yield has slightly improved, same yield, more or less same, low yield, better yield with manual | Yield mixed reviews |  | Processes |
| Far-flung distance, Van is unable to go, Scattered population, Security Issues, No point of perfect site if van can't reach, 70% inaccessible sites KPK, Location on mountain top, Narrow streets | Geographic/Access Issues | **Operational and contextual challenges** | Infrastructure |
| AI causes more area repetition over time, Repetition decreases options, Limited ability of AI, same locations | Technical Issues |  | Technology |
| No surety of perfect location with AI, AI selected area had 2 houses, No shelter near exact AI, MATCH doesn't consider local context, Manual method knew local fit, AI lacks ground insight, Van size is big | Contextual Gaps |  | Process |
| Family didn't show up because of distance, Far areas participant walk 1.5KM, culture with females, hard terrain areas for participants, No communities in 5-10KM radius | Gender related or Cultural factors |  | Culture |
| Manual data is human intelligence, Know local context in manual selection, In manual we could tell if notables would cooperate, Manual camps ensured family presence for tracing, Autonomy in manual planning, Finding pts through contact, Identified new cases and areas through TPT, Finding patients by medicine intake tracing, Manual method was done on our convenience, Camp near participants if given choice, choosing a middle area with most participants manually | Local Knowledge | **Challenges of the staff** | People |
| Several pre-camps visit, Meeting with area notables, Notables help reduce TB stigma, Importance of community awareness, Venue arrangements before camp, DFS visits all sites to select the best | Pre-Camp Activities |  | Process |
| Double work due to digitalization, unable to complete tasks in recommended time, Entering same thing at multiple places | Increased Burden | **Organizational readiness** | Technology |
| Appreciating digital application, Digital data easy to access, everyone has digital data access at same time | Positive Perceptions of Digital Systems |  | Technology |
| Application internet/sync issues, Interface shuts down, Can't make amendments/summary reports in digital, Data uploading/stuck issues, Digital errors, Data lost, Complex use of app, Multiple digital systems inconvenience, Missing and duplicate data Tablet vs. web version inconsistencies, MATCH AI algorithm updates multiple location recommendations | Technical Challenges |  | Infrastructure |
| DHO factor, DFS concerns and other issues, DTC complains, DTC not involved, Resistance to trying something new | Role of Stakeholders | **Stakeholder engagement** | People |
| MATCH-AI understanding, Unawareness on how MATCH works, Decisions taken at federal are applied uniformly without context, trickle down of information, power dynamics | System understanding | **across hierarchy** | People |
| Initial training but no refreshers, Not much training, Need for capacity building, Request for training/refreshers, Refreshers to discuss field challenges, Refreshers to stay on same page | Orientations and refreshers |  | Goal |

### Ethical Considerations

Ethical approval was obtained from the Health Services Academy, Islamabad, Pakistan (7– 82/IERC-HSA/2022–52) and from the Common Management Unit for TB, HIV and Malaria, Ministry of Health Services, Regulation and Coordination, Islamabad, Pakistan (CMUE-ERC- 26). Participation was voluntary, with informed consent, no incentives, and confidentiality was ensured through anonymization and secure data handling.

### Results Demographics

The study sample comprised of 42 participants from multiple SRs of TB program across Pakistan. Table 1 summarizes the selection of study participants across different provinces (Federal, Punjab, Sindh, KPK, and Balochistan). Most participants are male and hold field- and program-level positions such as District Field Supervisors, Project Coordinators, Regional Coordinators, and Program Managers. Experience levels vary widely, ranging from 6 months to 15 years, indicating a mix of early-career and highly experienced staff involved in the program. (see supplementary file 3)

### Theme 1: MATCH-AI as a useful tool

Most participants spoke positively about MATCH-AI, describing it as an important step toward using technology in selecting locations for TB screening camps. Before its introduction, camp site selection largely depended on local knowledge, stakeholder consultations, previous camp experiences, and historical TB records. Staff described how MATCH-AI reduced the need to manually search TB registers or rely heavily on doctors and local leaders to identify camp locations. Instead, the system automatically generated a list of sites, including some that were geographically distant or previously unknown to the teams. Although reaching these locations sometimes required additional effort, participants considered it worthwhile when camps conducted in these areas identified TB cases that may otherwise have been missed. ‘A project coordinator mentioned that: *‘Despite the distance, we had a case where four to five TB-positive individuals were identified in the same household in a remote area of Faisalabad. MATCH-AI marked that same location again, and the rider along with the DFS made the effort to visit it*.

#### Without this system, we might have missed these cases altogether.’ (E8, FGD)

Participants also felt that MATCH-AI introduced a more structured and transparent process by generating a list of potential camp locations based on data. Previously, conducting a camp could be influenced by personal preferences or external pressure from local authorities and public officials but now, they viewed MATCH-AI as a tool that improved transparency by reducing personal bias in selecting camp locations and helping them manage external pressure. One participant said: *‘Government officials also direct us to work in certain areas. For example, DTCs and DHOs sometimes ask us to hold camps where they are visiting. In such cases, we face pressure and usually rely on the 10% flexibility in our plan. However, since we have already shared our work plan, we mostly adhere to it. We also try to convince both political figures and public sector officers.’ – (D3, Balochistan)*

Despite its perceived benefits, participants had mixed views regarding the effectiveness of MATCH-AI in improving TB yield. A few of the participants believed that the yield has improved with the AI tool, while many others observed no differences between the two or even better TB yield with manual site selection method.

*“When we are getting an identified site with MATCH-AI, then definitely the yield gets good.” – (B5, Balochistan)*

*“Yield is also the same; it’s 1.5–1.6 with MATCH-AI and with manual it was 1.4–1.5.” – (B2, KPK)*

*“We are getting more cases manually compared to MATCH-AI.” – (A7, Balochistan)*

### Theme 2: Operational and contextual challenges

This theme highlights several interrelated issues and contextual factors affecting the suitability and usability of AI-generated location selection.

***Geographic/ Access Issues:*** A key limitation identified especially in the KPK region was the inaccessibility of locations identified by MATCH-AI. Participants shared that MATCH-AI sometimes suggested locations such as mountains, snow-covered areas, lakesides, or even areas outside their allocated districts. *“They gave me locations like Naran Kaghan, Babusar, Jheel Saif-ul-Malook. There’s 3 feet of snow. How will I get there?” – A1, KPK*

The system lacked local knowledge and did not acknowledge the security risks present in certain regions of Pakistan, particularly in Balochistan, and generated locations of high-risk security areas. *“The issue is that it does not consider security.” – (A7, Balochistan)*.

***Contextual Gaps*** Field staff reported that AI locations didn’t always translate into practical locations, with some sites having very few households or lacking basic accessibility. In contrast, manual selection was seen as more reliable due to its grounding in local knowledge and understanding of on-the-ground conditions for example the roads were narrow and the big sized van couldn’t pass through to reach the locations suggested by the AI.

***Technical Issues:*** Some participants mentioned that the ACF teams often revisited the same locations without finding any new cases suggested by the AI, which not only wasted resources but also missed out on opportunities to explore newer areas. *“If it’s in the same place again and again, it wastes our energy, the doctor’s, and affects the community too”. – (D2, Punjab)*

***Cultural Factors***: In culturally conservative areas particularly in Khyber Pakhtunkhwa, gender norms significantly influenced accessibility. Field staff mentioned that female patients were often reluctant or even forbidden to travel long distances by their male relatives, especially if the camp location is public or lacks privacy. *“Women here cover themselves. If the van is parked 5 km away, in the market, they won’t go. They’ll say, ‘Even if I die, I won’t go.’” – (A10, KPK)*

Sometimes there was no population density or camps were not accessible to people especially the poor, elderly, or women without transport. *“TB is a disease of the poor. Do you think someone will walk 5 km just to reach the camp? People ride bikes for food, not TB check-ups.” – A10, KPK*

Most mobile camps lack private spaces, making women uncomfortable with testing or discussing health issues in public, mixed-gender settings. Field staff emphasized that with manual planning, they could ensure the van was parked within the community or near homes, ensuring women felt safe and respected*. “I always manage to bring females to camps because I know the area. AI doesn’t consider that.” – A10, KPK*

### Theme 3: Challenges of the staff

Field staff consistently emphasized that while AI-based systems like MATCH-AI can identify distant high-burden geographic areas, the effectiveness of TB screening depends heavily on how these outputs are interpreted and implemented on the ground.

A central aspect of this contextual decision-making was the autonomy field staff used to exercise in planning screening activities. Drawing on prior experience in similar settings, staff strategically determined where to position camps, how to organize contact tracing, and how to sequence interventions such as TB Preventive Treatment (TPT). One participant explained: *‘’During camp, we had multiple responsibilities: we had to give TPT, do contact tracing, and cover the area too. AI compromises all of that. With manual planning, based on field experience, I could decide: I’ll bring the van here, I’ll conduct TPT here, I’ll cover so and so contacts here and there, and so on. It was very strategically planned.’’ – A11, KPK*

Beyond logistical planning, community engagement emerged as a critical determinant of screening success. Pre-camp visits and meetings with local notables were repeatedly described as essential steps in mobilizing participation, particularly in settings where TB-related stigma remains a barrier to care-seeking. Notables functioned as trusted intermediaries who helped bridge the gap between implementers and community members, thereby increasing acceptance of screening activities. As one participant noted: *“Participation is directly linked to notable activities. AI has nothing to do with participation.” – E8, FGD*.

Field staff described pre-camp engagement as a structured process involving multiple visits to the lists of sites provided, discussions with community leaders, coordination with local healthcare providers, and logistical preparation. These interactions were not merely administrative but were central to building trust and ensuring that screening camps were accessible. In contexts where stigma around TB persists, such trust-building efforts were seen as more influential than the technical identification of high-burden areas alone. However, this reliance on field-based adaptation also introduced significant operational challenges. Staff reported increased travel distances, repeated site visits, and additional coordination responsibilities, particularly after AI-generated site selection expanded the geographic scope of fieldwork. One supervisor highlighted the growing workload: *“It is just that the load has become more as first we used to stay at 20KM but now we have to travel up to 40KM. We also have to do compulsory 3-4 visits.” – A6, Punjab*

This also carried financial and logistical implications, including fuel costs and time constraints that were often not formally compensated. As another participant explained, extended travel and repeated visits often consumed entire working days, delaying other essential tasks and contributing to fatigue. *"Personally, I never had issues with fuel costs, but other DFSs are very concerned about these expenses. They say that we have to do 4 visits in such far areas, the weather is so hot, and additionally, we spend our own fuel. The time is limited because the work burden is much. As it is becoming digitalized, we work in both soft and hard forms. So, time is a big issue. If we go far away, our whole day goes into this and other tasks stay pending." -A6, Punjab*

Across all sites, managers and coordinators reinforced that successful TB screening was not determined solely by identifying the right locations, but community participation was directly proportional to mobilization activities. In this sense, local knowledge and community mobilization were not supplementary to technology but essential to its successful implementation in practice.

### Theme 4: Organizational readiness

The electronic Case-Based Surveillance (eCBS) system was introduced in parallel of the MATCH-AI implementation to enable real-time, location-based data collection from field camps, which would continuously feed into the AI model to generate updated and more accurate TB sites predictions. Since the AI model depends on timely field updates, delays or gaps in data entry directly affected its ability to generate refined and efficient site recommendations. Field staff reported persistent issues such as unstable internet connectivity, system lag, and frequent server failures, particularly in remote areas where connectivity was either weak or unavailable. As one participant explained: *“Sometimes our maps are down. Sometimes our server is down*.

*Sometimes, the events that we make disappear. We can’t find them. Sometimes, when we go there, there is no internet access… without internet accessibility, there are no entries. Sometimes we have issues in syncing.”- A9, Sindh*

As a result, many workers reverted to paper-based recording in the field and later duplicated entries into the digital system when connectivity improved. This dual documentation process increased workload, created repetitive data tasks, and contributed to frustration among staff. *“We make sheets as fast as we can, and we enter into eCBS. After entry, you have to come to your own territory. Sometimes, when you camp 10–15 km further, you mostly have to come back to the city.” – A5, Punjab*

This led to field staff frequently experienced strain in balancing digitalized planning systems with extensive on-ground responsibilities. This was also one of the reasons behind technical issues like repetition of locations as mentioned during the conversation in FGD with data managers: ‘’*Sometimes issues like repetition come up, so we guide them reminding them that they already have 30 MATCH recommendations and need to choose from these multiple options. The purpose of introducing eCBS was that when the data from camp will be updated in real time, the MATCH algorithm will pick it and generate new locations but the process wasn’t smooth because of various technical issues at our end and then issues at the user end like duplication of records and digital illiteracy.’’ -F8, FGD*

### Theme 5: Stakeholder engagement across hierarchy

The implementation of MATCH-AI revealed varying levels of engagement and alignment among key stakeholders involved in TB screening activities. In the manual system, district-level actors such as District Health Officers (DHOs) and DTCs played a direct role in selecting sites and approving camp locations, which ensured their active involvement in decision-making. However, with the introduction of AI-supported site selection, their roles became less defined, leading to inconsistent levels of engagement across districts. In some areas, district authorities remained actively involved, while in others they were largely disengaged from the process, contributing to reduced coherence in implementation. For example, in one district, a DTC declined to participate in an interview, stating he was not interested and was too busy, while in another district, a DTC who was interviewed said, *‘With AI, how sites are detected? This is the first time we are doing it through AI. We don’t know how big of a role AI has. In foreign countries, it’s different; in our peripheries, it is different. In foreign countries, there are more educated people who understand things better and reach hospitals and facilities, but here, we don’t have that. TB patients are dying at home and they don’t know. So, these kinds of patients, how will you get them through AI?’ -C3, Balochistan*

This shift also exposed a broader challenge of limited understanding of how the AI functions among different stakeholder levels. While senior managers demonstrated partial awareness of the purpose of MATCH-AI, understanding declined progressively at regional and field levels. Field supervisors in particular reported uncertainty regarding how locations were selected or how the system processed data. As one participant noted, *“We don’t know which locations are selected by AI or on what basis. We just know that they have all the data.” – B1, KPK*

This lack of clarity was further compounded by communication gaps between administrative levels. Changes in system processes or documentation were often communicated at higher levels but did not consistently reach field staff in a timely or structured manner. As one participant explained, *“When changes happen, the people in higher positions know, but the people on the ground don’t necessarily know.” – A3, KPK*

Across interviews, stakeholders highlighted that limited orientation and training on MATCH-AI reduced confidence and ownership among field staff. While some managerial level participants acknowledged attempts to explain the intervention, they also noted that varying levels of education and digital literacy constrained full comprehension of AI-based decision-making processes. As project coordinators reflected in the FGD, *“They (DFS) were completely unaware. Without proper orientation, understanding cannot trickle down. Even when we didn’t have formal orientation, we tried to understand and explained it multiple times through presentations. But the challenge was their limited technological background, they could not fully grasp how AI works.’ – E8, FGD*

Beyond technical deployment, the effectiveness of AI-supported TB screening is strongly shaped by stakeholder alignment, communication flow, and shared understanding of the intervention.

Where these elements were weak, the integration of AI into routine field operations remained partial and uneven.

## Discussion

This study explored the perceptions implementing the use of an AI-assisted software for the site selection of chest camps conducted for active case finding of TB cases in Pakistan. While there is emerging evidence showing that AI supported technologies in health sciences are efficient, it is important to understand the contextual realities and on the ground support needed to make it work in pragmatic settings.^14^ This study has looked at various operational factors from a user’s perspective including tool’s accuracy, the prerequisites required for its successful adoption, alignment between technical systems and social conditions, community involvement and optimal orientation of the implementing staff. Using STS framework, the findings demonstrate that technology and social factors are closely linked and AI can substantially strengthen TB outreach and screening programs when integrated with local experience and knowledge, health system preparedness and contextual adaptation.^11,12^

A key findings that emerged is the positive perception of MATCH-AI as an evidence-based and non-biased approach to camp site selection. Across all districts, participants consistently emphasized that choosing the right location is critical to achieving high TB yields particularly because ACF is a resource-intensive activity.^3,15^ These findings are consistent with existing literature demonstrating that AI-supported and digital health technologies can improve efficiency and reduce the human workload within TB programs and other healthcare settings.^16,17^ Further to this, participants believed that MATCH-AI helped reduce selection bias as local leaders often influence the selection of a camp location for personal or political reasons. This supports the field staff manage external pressures exerted by community and reduced selection bias.^9^ Although not consistently reported by all participants, some areas selected by MATCH-AI noted higher camp yields, by identifying new locations that were overlooked in manual site selection and uncovered “sites never known to have existed.” Thus, the use of MATCH-AI can support more targeted ACF interventions by identifying new clusters of high transmission which in a high TB burden setting like Pakistan can significantly improve efficiency of ACF programs.

Although MATCH-AI was generally viewed as a promising tool to improve the efficiency of ACF approach, substantial gaps were highlighted between AI generated outputs and cultural context, ground realities and operational feasibility providing potential explanations for the low and inconsistent adherence to AI-recommended sites observed in the SPOT-TB trial.^5^ Sites were often inaccessible due to terrain, security, poor infrastructure or transportation barriers — consistent with existing literature. ^9,18^ Our findings provide further evidence that use of AI may not always translate into effective field implementation because of existing field realities like logistical constraints in reaching recommended locations. The participants of our study valued MATCH-AI recommendations but consistently suggested that the recommendations provided by AI should be supplemented by their own knowledge based on field experiences and local context. Thus, while AI provides non-biased objective selection of sites, field staff can provide contextual awareness and operational flexibility that cannot be replicated by AI alone.^17^ As a result, manual camp selection is preferable when community engagement is needed to obtain local permissions and identify appropriate locations for conducting camps. This builds community trust and ownership which is essential for TB response success. ^19–21^ Repeated recommendation of certain locations was also reported as another major challenge, and finding highlights the importance of continuous data updating and feedback into the MATCH-AI system, which otherwise loses its effectiveness.

Another key finding that emerged out of this study is the organizational readiness and the preparedness of the implementing staff to take up a new technology such as MATCH-AI. Field staff across all geographies, reported multiple technical and capacity related challenges that led to an ineffective implementation of the MATCH-AI supported approach. Other than human capacity challenges, various technical issues such as poor internet connectivity, data synchronization issues, and operational ineffectiveness of the digital data system were identified by the participants. Although the eCBS application had been introduced as part of the program’s broader digitalization efforts before MATCH-AI, the two interventions were implemented within a relatively short timeframe and became closely integrated, as MATCH-AI relied on digital data collected through eCBS. Consequently, participants’ experiences with digitalization and MATCH-AI may have overlapped, making it challenging to distinguish perceptions attributable specifically to MATCH-AI. These observations are consistent with previous research that shows that switching from manual data systems to digital transitions without proper training and preparing the staff adequately can lead to work overload and user’s trust in AI-supported systems.^6,9,22^ When MATCH-AI was introduced, orientation sessions were conducted for each implementing organization to familiarize staff with the system. However, these sessions did not appear to generate a comprehensive and shared understanding of the intervention across all levels of implementation. Despite being most directly involved in implementing AI-recommended site selections, field staff often reported limited knowledge of how recommendations were generated or the principles underpinning the approach. Instead, implementation was frequently described as a process of following instructions communicated through supervisory channels. These findings suggest that hierarchical decision-making structures may have constrained opportunities for frontline engagement with, and critical appraisal of, the intervention, potentially affecting ownership, trust, and adherence during implementation.^7,9^ Participants consistently highlighted the lack of practical guidance, and the need for ongoing capacity building. Similar findings have been reported in studies, where inadequate training is associated with poor adoption of digital health innovations thereby resulting in overall ineffectiveness. ^8^

The study had some limitations. Given the context-specific nature of MATCH-AI and its implementation within the ACF setting, these findings should not be generalized to AI applications in healthcare more broadly. Rather, they provide insights into the implementation of this particular tool within this specific setting. Female participants were fairly underrepresented, reflecting the gender composition of the ACF workforce within the programme rather than intentional participant selection. The study focused on programme implementers and did not include patients or policymakers. In addition, the cross-sectional design meant that a before-and-after comparison of the AI intervention was not conducted.

Despite these limitations this study has provided valuable insights into stakeholder perceptions regarding the use of MATCH-AI for TB camp site selection in Pakistan, compared with traditional human based approaches. Our results demonstrated a fair acceptance of MATCH-AI among participants as an innovative and more systematic and evidence-based approach which reduces subjective selection of sites for holding screening camps thus eliminating selection bias. However, despite advantages, there exists a critical gap between algorithmic recommendations and ground realities suggesting that human experience and expertise remain crucial in decision making. These findings are best interpreted using the STS theory, which suggests that the best utility of AI-assisted tools such as MATCH-AI as TB depends on models that combine technology with contextual knowledge, community engagement, and field implementation experience. Finally, TB programs should strengthen data systems and provide training to support AI adoption.

## Supporting information

Coreq Checklist

Topic Guides

Consent Form

Demographic Characteristics

STS Interpretation

## Data Availability

All data produced in the present work are contained in the manuscript.

## Author Contributions

AL acquired the funding. FE, SMAZ, AM and AL conceptualized the study and wrote the protocol. AS, AF and SMAZ contributed to the literature search. SMAZ, AM, AS and AF designed the topic guides. AL, AM and FE conducted stakeholder consultation. NN and WN handled the filed coordination and took field notes. AM and AS conducted the interviews. AS and AF transcribed, translated the audios, accessed the data and were involved in the analysis. AS, AF, AM, AL and FE finalized the themes. AS wrote the first draft. TER and FE reviewed and the manuscript. AL and FE were responsible for the decision to submit the manuscript. All authors read, gave comments and approved the final manuscript.

## Data Availability

All relevant data are included in the paper and supplementary files. The study’s audio files and transcripts contain sensitive and personal information about the participants, and cannot be shared under current approvals to maintain confidentiality.

## Acknowledgements

We thank the National TB Control Program Pakistan for supporting and leading the evaluation of MATCH-AI for optimizing placement of TB screening activities in Pakistan. MATCH-AI was co-developed by KIT Royal Tropical Institute and EPCON, whose technical collaboration supported implementation of the intervention evaluated in this study.

We thank Mercy Corps for leading study implementation and the Center for Global Public Health for supporting study design, data analysis, and independent evaluation. We are grateful to all implementing partners: Association for Community Development (ACD), Bridge Consultants Foundation (BCF), Marie Adelaide Leprosy Centre (MALC), Association for Social Development (ASD), Greenstar Social Marketing (GSM), and Strengthening Participatory Organization (SPO) as well as the Mercy Corps Project Implementation Unit (MC-PIU), Mercy Corps Quetta (MC-Quetta), and Mercy Corps Hyderabad Sindh (MC-Hyderabad Sindh) offices and field teams for their contributions to implementation of the stepped-wedge trial and community screening activities.

Finally, we thank Guy Stallworthy and Puneet Dewan from the Bill & Melinda Gates Foundation for their support and technical guidance throughout the project.

## Research in Context Evidence before this study

We reviewed existing literature from 2019 till 2025 using PubMed/MEDLINE and Google Scholar. Additional grey literature and institutional publications were identified through targeted searches of the institutional websites and ResearchGate. Literature search was done in English language and the search terms included combinations of: "artificial intelligence, “tuberculosis," "active case finding," "AI interventions in healthcare/TB", "AI implementation barriers," "low- and middle-income countries," and "sociotechnical systems." Studies were included if they addressed AI or digital technology use in TB screening or public health surveillance, implementation barriers and facilitators in LMIC contexts, or community engagement in TB programs

This qualitative study was conducted to compliment the on-going SPOT TB trial which used MATCH-AI, an artificial intelligence tool to recommend where TB camps should be held to screen people. Whether such tools work well once they meet the field is a different question from whether they perform well on paper. Studies of AI-based chest X-ray reading tools for TB in India and other high-burden countries have shown that even accurate algorithms can run into real-world trouble: patchy data, unreliable connectivity, and field staff who weren’t given enough training or explanation to trust the system. Broader research on bringing AI into healthcare in lower-income settings echoes this that strong performance in testing does not guarantee strong performance in everyday operations, and adoption often hinges as much on organizational readiness and staff confidence as on the algorithm itself. These findings complement to the SPOT TB trial and an understanding of why: how field staff and stakeholders actually experienced the tool and what led them to adhere to its recommendations or fall back on their own judgement.

## Added value of this study

While most research on AI in TB has focused on diagnostic applications, particularly automated chest X-ray interpretation, this study examines a comparatively underexplored application: the use of AI to support decisions about where to locate active case-finding screening camps.

Embedded within a multicenter implementation trial across 14 districts in four provinces, this qualitative study draws on perspectives from frontline field staff, supervisors, and program managers to provide a comprehensive understanding of how AI-supported planning is implemented in routine practice. By integrating views across different health system levels and implementation contexts, the study advances understanding of the contextual, organizational, and operational factors that shape the adoption and use of AI-enabled decision-support tools. It provides a human-centered explanation of implementation processes that complements the evidence from SPOT TB trial and helps explain how context influences the effectiveness of AI- supported public health interventions.

## Implications of all the available evidence

Together, the SPOT TB trial and this study suggest that AI-guided targeting can improve TB detection, but only when implementation is supported by the health system and local context. The available evidence indicates that AI-based planning tools should be used to support, rather than replace, local decision making, with recommendations treated as a starting point that staff can adapt based on contextual knowledge. These findings also highlight the importance of regularly updated data, training that reaches frontline field staff as well as managers, and reliable digital infrastructure to support implementation. Future research should evaluate hybrid human– AI workflows, assess models that use continuously updated data, and embed implementation research within AI-health trials to understand how contextual factors, including gender and community-level access barriers, influence effectiveness alongside trial outcomes.

## Supplementary Materials Available in a separate file Use of Generative AI

The authors used generative AI only for language and grammar editing to improve readability during manuscript preparation. The authors reviewed and edited the content as needed and take full responsibility for the content of the publication.

