## Supplementary material for "Geographical targeting of active case finding for tuberculosis in Pakistan using artificial intelligence software: a qualitative study embedded within the SPOT TB trial": Topic Guides

**Topic Guide for RC, DFS, PM**

**Objective/Purpose:**

This study is planned with the aim “to explore the barriers and facilitators in existing and new (MATCH AI) practices for camp planning and uptake’’.

The study will explore:

- The perspectives and experiences of healthcare workers while using MATCH-AI to identify the locations of ACF camps with focus on barriers and facilitators of uptake of intervention.
- Investigate how using AI has affected staff efficiency in terms of ACF planning, workloads, processes and data management, relative to the conventional approach.

**Introduction:**

Assalam o alaikum. My name is Alina and I am a qualitative researcher at CGPH. We are working as a third person evaluation party for this MATCH AI intervention. Thank you for participating in this important study. Today, we’ll be exploring your experiences and perspectives on using the MATCH AI software for site selection in ACF camp planning in Pakistan. Artificial intelligence uses data to make different types of predictions. In this project we used AI to select locations of ACF camps.

The goal of this session is to understand the challenges and barriers you’ve encountered when using MATCH AI, as well as compare this approach with the traditional methods currently in use.

There has been limited study of how AI supports these camps and what practically works (does not work) in the field setting. Your insights will help us better understand the advantages and limitations of both the MATCH AI method and the conventional approach. We’ll also look at how the revised approach has impacted your efficiency in planning, data management, and overall workload.

This session is designed to be an open and informal conversation, so please feel free to share your honest thoughts and experiences. There are no right or wrong answers, and your input will be crucial in helping us improve the process for healthcare workers and the planning of future ACF camps. We will go with the questions one by one and maintain a flow. Your names will not be recorded to maintain confidentiality.

We appreciate your time and the valuable feedback you are providing to shape the future of this process.

**Questions:**

**Demographic Information:** Gender, District, Role, Years of experience in your current role, Years of experience with ACF, Experience with MATCH AI.

As we know, ACF is a strategy used to increase case detection in a scientific and systematic screening for undiagnosed TB or other infectious diseases outside of health facilities. So, with that being said,

**Role and Responsibilities**

1. Can you describe your role in the organization and how it relates to ACF?

**Importance of Location**

We know that there are two approaches with location: one is the method you were using prior to Match AI, where you would perhaps go out in the field, operate manually by using gained knowledge, experiences, and analysis of historic data provided by the field staff or other authoritative figures. However, with the introduction of MATCH-AI, you now get pre-decided locations from the office. With locations of the campsite in mind:

1. How important do you think the location of the campsite is, and why?
2. What factors do you consider for your selection?
3. What was your role in the previous method? What is your role today?
4. How does MATCH-AI work? How is the match AI different from the previous old **way**?
5. Did you see a difference in your **role** after the introduction of MATCH AI? Do you believe AI has made management easier or more difficult for you?
6. Do you think this new method of MATCH-AI considers local context (your situation/ setting) when selecting sites?
7. Are there difficulties in locating different camp sites? If so, what kind of difficulties?

**Efficiency and Workload**

1. What was the initial response of your team members and supervisors in implementing MATCH-AI?
2. Do you think AI has made conducting camps easier or difficult for you?

**Barriers and facilitators**

Van teams had option of conducting camps without MATCH-AI as well

1. What made you decide that a camp should be done using MATCH-AI?
2. What made you decide not use locations of MATCH-AI?
3. Did you encounter difficulties in locating different camp sites selected by MATCH-AI? If so, what kind of difficulties?

**Improving Camp Yields and Performance**

Since you have been implementing MATCH AI for a while now.

1. In your opinion, what activities or strategies can help improve the yield and performance of the camps conducted through AI spot identification? What factors make a camp more successful in finding cases?
2. After MATCH AI: What is the turnover of people in camps on average? How many cases per camp do you get on average?
3. Notable meetings: who does it, how is it done? when, how, why… the turnover?

**Digital Data Usefulness**

Recently, you have been using digital data management tools like eCBS, while previously all data was managed on paper.

1. Which medium do you find better to use: paper-based or digital? Why or why not?
2. Is capturing digital data easy and/or useful?
3. What challenges do you face with the usage of digital tools?
4. Did you get any training or refreshers on using digital tools? How was it? Feedback.

**Staff Training**

1. What kind of additional support and/or training do you need?

**Future Use and Adaptation**

1. Would you like to incorporate long-term adoption of MATCH AI? Why or why not?

**Further Information**

1. Is there anything/something you would like to share that we haven’t asked?

**Additional Notes:**

- **Context and Background Information:**

The primary focus of this session is to understand the challenges healthcare workers face when using the MATCH AI technique for ACF camp site selection in Pakistan. We also aim to compare this AI-based approach with the conventional method. Understanding your experience will provide valuable insights into the practical application of both methods and help identify areas for improvement.

- **What MATCH-AI study adds:**

SPOT-TB is a pragmatic stepped wedge cluster randomized trial that will evaluate whether a targeted approach towards ACF, supported by artificial-intelligence software (MATCH-AI), can increase yields of TB cases detected. The study is being conducted on a national scale in Pakistan and is embedded within the routine operations of the implementing partners of the National TB Program. The study will provide empirical evidence for the potential benefit of targeting ACF interventions in a real-world setting, within a high-burden TB country.

- **Specific Instructions:**

Participants are encouraged to share both positive and negative experiences with the MATCH AI system. We are interested in how the system impacts your daily operations, your workflows, and overall staff efficiency. Please be candid and share as much detail as possible, as your input will guide future improvements.

- **Managing Responses and Encouraging Discussion:**

It is important to ensure that all participants feel comfortable expressing their opinions. If a participant is hesitant, gently prompt them with follow-up questions to draw out deeper insights. For example, ask about specific experiences or challenges. Ensure everyone has an opportunity to speak by keeping track of responses and directing questions to quieter participants.

- **Managing Time:**

Aim to stick to the suggested timings for each section to ensure the session remains focused. If a discussion is going off-topic or too detailed, kindly steer it back to the specific questions or themes.

**Closing Remarks:**

Thank you so much for your time and valuable contributions today. Your feedback is extremely helpful in improving the MATCH AI system and enhancing the ACF camp planning process. We will analyze the information gathered from all participants and use it to identify key areas for improvement in both the AI technique and the traditional camp site selection approach. We’ll follow up with a summary of the findings, and any significant changes or next steps will be communicated to you in due course. If you have any further thoughts after today’s session, feel free to reach out.

**Timing/Duration:**

Estimated Total Duration: 45- 60 minutes

**Topic Guide for DTC**

**Objective/Purpose:**

This study is planned with the aim “to explore the barriers and facilitators in existing and new (MATCH AI) practices for camp planning and uptake’’.

The study will explore:

- The perspectives and experiences of healthcare workers while using MATCH-AI to identify the locations of ACF camps with focus on barriers and facilitators of uptake of intervention.
- Investigate how using AI has affected staff efficiency in terms of ACF planning, workloads, processes and data management, relative to the conventional approach.

**Introduction:**

Assalam o alaikum. My name is Alina and I am a qualitative researcher at CGPH. We are working as a third person evaluation party for this MATCH AI intervention. Thank you for participating in this important study. Today, we’ll be exploring your experiences and perspectives on using the MATCH AI software for site selection in ACF camp planning in Pakistan. Artificial intelligence uses data to make different types of predictions. In this project we used AI to select locations of ACF camps.

The goal of this session is to understand the challenges and barriers you’ve encountered when using MATCH AI, as well as compare this approach with the traditional methods currently in use.

There has been limited study of how AI supports these camps and what practically works (does not work) in the field setting. Your insights will help us better understand the advantages and limitations of both the MATCH AI method and the conventional approach. We’ll also look at how the revised approach has impacted your efficiency in planning, data management, and overall workload.

This session is designed to be an open and informal conversation, so please feel free to share your honest thoughts and experiences. There are no right or wrong answers, and your input will be crucial in helping us improve the process for healthcare workers and the planning of future ACF camps. We will go with the questions one by one and maintain a flow. Your names will not be recorded to maintain confidentiality.

We appreciate your time and the valuable feedback you are providing to shape the future of this process.

**Questions:**

**Demographic Information:** Gender, District, Role, Years of experience in your current role, Years of experience with ACF, Experience with MATCH AI.

As we know, ACF is a strategy used to increase case detection in a scientific and systematic screening for undiagnosed TB or other infectious diseases outside of health facilities. So, with that being said,

**Role and Responsibilities**

- Can you describe your role and how it relates to camp selection for identification of TB cases?
- How do you give approval of the camp plan? What are the steps you take to finalize a plan?

**Importance of Location**

We know that there are two approaches with location: one is the method you were using prior to Match AI, where with your guidance, DFS would consider the tb03 form or perhaps go out in the field, operate manually by using gained knowledge, experiences, and analysis of historic data provided by the field staff or other authoritative figures. or some political influence

- So do you have to listen to other influential people in your district or province while finalizing the camp plan?
- How important do you think the location of the campsite is, and why?
- How do you guide the staff on selecting the site?

However, with the introduction of MATCH-AI, you now get pre-decided locations from the office. With locations of the campsite in mind:

- What was your role in the previous method? What is your role today?
- How does MATCH-AI work? How is the match AI different from the previous old way?
- Did you see a difference in your workload after the introduction of MATCH AI? Do you believe AI has made management easier or more difficult for you?
- Do you think this new method of MATCH-AI considers local context (your situation/ setting) when selecting sites?
- Are there difficulties in locating different camp sites? If so, what kind of difficulties?

**Efficiency and Workload**

- What was the initial response of overall people involved in camp selection when MATCH-AI was introduced?
- Do you think AI has made conducting camps easier or difficult for you?

**Barriers and facilitators**

You get recommendations every month but sometimes the recommendations are not considered/ followed.

- What are the reasons behind this in your opinion?

**Improving Camp Yields and Performance**

Approximately how much time has it been since you are implementing MATCH AI?

- In your opinion, what activities or strategies can help improve the yield and performance of the camps conducted through AI spot identification? What factors make a camp more successful in finding cases?
- Do you have any idea if the turnover is the same or has increased or decreased?
- In your district, have you noticed the difference in the number of camps? Do you think these camps are adequate?

**Future Use and Adaptation**

- Would you like to incorporate long-term adoption of MATCH AI? Why or why not?

**Further Information**

- Is there anything/something you would like to share that we haven’t asked?

**Additional Notes:**

- **Context and Background Information:**

The primary focus of this session is to understand the challenges healthcare workers face when using the MATCH AI technique for ACF camp site selection in Pakistan. We also aim to compare this AI-based approach with the conventional method. Understanding your experience will provide valuable insights into the practical application of both methods and help identify areas for improvement.

- **What MATCH-AI study adds:**

SPOT-TB is a pragmatic stepped wedge cluster randomized trial that will evaluate whether a targeted approach towards ACF, supported by artificial-intelligence software (MATCH-AI), can increase yields of TB cases detected. The study is being conducted on a national scale in Pakistan and is embedded within the routine operations of the implementing partners of the National TB Program. The study will provide empirical evidence for the potential benefit of targeting ACF interventions in a real-world setting, within a high-burden TB country.

- **Specific Instructions:**

Participants are encouraged to share both positive and negative experiences with the MATCH AI system. We are interested in how the system impacts your daily operations, your workflows, and overall staff efficiency. Please be candid and share as much detail as possible, as your input will guide future improvements.

- **Managing Responses and Encouraging Discussion:**

It is important to ensure that all participants feel comfortable expressing their opinions. If a participant is hesitant, gently prompt them with follow-up questions to draw out deeper insights. For example, ask about specific experiences or challenges. Ensure everyone has an opportunity to speak by keeping track of responses and directing questions to quieter participants.

- **Managing Time:**

Aim to stick to the suggested timings for each section to ensure the session remains focused. If a discussion is going off-topic or too detailed, kindly steer it back to the specific questions or themes.

**Closing Remarks:**

Thank you so much for your time and valuable contributions today. Your feedback is extremely helpful in improving the MATCH AI system and enhancing the ACF camp planning process. We will analyze the information gathered from all participants and use it to identify key areas for improvement in both the AI technique and the traditional camp site selection approach. We’ll follow up with a summary of the findings, and any significant changes or next steps will be communicated to you in due course. If you have any further thoughts after today’s session, feel free to reach out.

**Timing/Duration:**

Estimated Total Duration: 45- 60 minutes

**Topic Guide for Focus Group Discussion with ACF Team**

**Objective/Purpose:**

This study is planned with the aim “to explore the barriers and facilitators in existing and new (MATCH AI) practices for camp planning and uptake’’.

The study will explore:

- The perspectives and experiences of healthcare workers while using MATCH-AI to identify the locations of ACF camps with focus on barriers and facilitators of uptake of intervention.
- Investigate how using AI has affected staff efficiency in terms of ACF planning, workloads, processes and data management, relative to the conventional approach.

**Target Audience:** Data Managers ACF

**Introduction:**

Assalam o alaikum. My name is Alina and I am a qualitative researcher at CGPH. We are working as a third person evaluation party for this MATCH AI intervention.

Welcome, and thank you all for joining this focus group discussion. We deeply appreciate the critical role you play in managing, interpreting, and sharing data, especially when it comes to using MATCH AI for identifying and prioritizing locations for ACF activities.

Today’s discussion aims to explore your day-to-day experiences, challenges, and perspectives related to data use - both digital and paper-based - in TB camp planning and implementation. We want to better understand how you assess data quality, how MATCH AI supports your decision-making, and what gaps or improvements you feel are needed. Your insights will be invaluable in strengthening data-driven strategies and ensuring better alignment between AI-generated recommendations and on-ground realities. We encourage open sharing, there are no right or wrong answers. We’re here to listen, learn from your expertise, and understand how data management practices can be enhanced to support effective TB screening efforts.

**Focus Group Discussion Questions**

**1. Person’s Role in the Organization**

- Can you briefly describe your role and responsibilities in your organization, particularly in relation to data management and reporting?
- How are you involved in reviewing or disseminating MATCH AI data?
- Who do you regularly collaborate with when using this data?

**2. Perspective on ACF and Its Utility**

- What is your understanding of the Active Case Finding approach in your current work?
- From your perspective as a data manager, how effective is ACF in identifying TB cases or improving service delivery?
- How do you use the SRs feedback monthly? Do you think the current ACF strategy is data-driven enough? Why or why not?

**3. Data Digitalization vs. Paper-Based Data Collection**

- What types of data collection methods are currently used in your organization (paper-based, digital, or hybrid)?
- How has the shift (if any) from paper-based to digital data collection affected your workflow?
- What challenges have you encountered in either method?
- What feedback do you have on ECBS and how are you tackling it?

**4. Merits and Demerits of Each Approach**

- Based on your experience, what are the key advantages of digital data collection (e.g., speed, accuracy, accessibility)?
- What are the limitations or risks associated with digital tools?
- In what situations (if any) do you still prefer paper-based data collection?

**5. Using Data for Camp Selection**

- How do you currently use MATCH AI to determine potential locations for TB screening camps?
- In your view, how can MATCH AI or other data sets be used more effectively for selecting locations for ACF camps?
- What filters, variables, or patterns do you prioritize…. Are there specific indicators or trends you look at to decide the best sites?
- Can you describe a situation where data directly influenced camp placement decisions?
- How well does MATCH AI align with ground realities reported by SRs or field teams?

**7. Accuracy and Precision of Camp Location Recommendations**

- In your experience, how accurate and reliable are the MATCH AI-generated locations for ACF?
- What metrics or feedback mechanisms do you use to assess the accuracy of these suggestions?
- Are there gaps or discrepancies between AI-recommended locations and field realities?

**8. Quality Checks for Data**

- What steps or protocols do you follow to verify the quality and integrity of the data being collected and reported?
- Are there tools or dashboards in place to track data inconsistencies or errors?
- How often do you conduct data quality audits or validation exercises?

Thank you so much for your time and valuable contributions today. Your feedback is extremely helpful in improving the MATCH AI system and enhancing the ACF camp planning process. We will analyze the information gathered from all participants and use it to identify key areas for improvement in both the AI technique and the traditional camp site selection approach. We’ll follow up with a summary of the findings, and any significant changes or next steps will be communicated to you in due course. If you have any further thoughts after today’s session, feel free to reach out.

**Topic Guide for Focus Group Discussion with Project Coordinators**

**Objective/Purpose:**

This study is planned with the aim “to explore the barriers and facilitators in existing and new (MATCH AI) practices for camp planning and uptake’’.

The study will explore:

- The perspectives and experiences of healthcare workers while using MATCH-AI to identify the locations of ACF camps with focus on barriers and facilitators of uptake of intervention.
- Investigate how using AI has affected staff efficiency in terms of ACF planning, workloads, processes and data management, relative to the conventional approach.

**Introduction:**

Assalam o alaikum. My name is Alina and I am a qualitative researcher at CGPH. We are working as a third person evaluation party for this MATCH AI intervention. Thank you for participating in this important study. Today, we’ll be exploring your experiences and perspectives on using the MATCH AI software for site selection in ACF camp planning in Pakistan. Artificial intelligence uses data to make different types of predictions. In this project we used AI to select locations of ACF camps.

The goal of this session is to understand the challenges and barriers you’ve encountered when using MATCH AI, as well as compare this approach with the traditional methods currently in use.

There has been limited study of how AI supports these camps and what practically works (does not work) in the field setting. Your insights will help us better understand the advantages and limitations of both the MATCH AI method and the conventional approach. We’ll also look at how the revised approach has impacted your efficiency in planning, data management, and overall workload.

This session is designed to be an open and informal conversation, so please feel free to share your honest thoughts and experiences. There are no right or wrong answers, and your input will be crucial in helping us improve the process for healthcare workers and the planning of future ACF camps. We will go with the questions one by one and maintain a flow. Your names will not be recorded to maintain confidentiality.

We appreciate your time and the valuable feedback you are providing to shape the future of this process.

**Questions:**

**Demographic Information:** Gender, District, Role, Years of experience in your current role, Years of experience with ACF, Experience with MATCH AI.

As we know, ACF is a strategy used to increase case detection in a scientific and systematic screening for undiagnosed TB or other infectious diseases outside of health facilities. So, with that being said,

**Role and Responsibilities**

2. Can you all one by one describe your role in the organization and how it relates to ACF?

**Importance of Location**

We know that there are two approaches with location: one is the method field teams were using prior to Match AI, where you would perhaps go out in the field, operate manually by using gained knowledge, experiences, and analysis of historic data provided by the field staff or other authoritative figures. However, with the introduction of MATCH-AI, you now get pre-decided locations from the office. Since you are overlooking all districts and managing the field staff. With locations of the campsite in mind:

8. How important do you think the location of the campsite is, and why?
9. What factors do field staff consider for selection?
10. Did they share about their previous role and the current after intervention?
11. How does MATCH-AI work? How is the match AI different from the previous old **way**?
12. Did you see a difference in your **role** after the introduction of MATCH AI? Do you believe AI has made management easier or more difficult for you?
13. Do you think this new method of MATCH-AI considers local context (your situation/ setting) when selecting sites?
14. Do you get feedback on difficulties in locating different camp sites? If so, what kind of difficulties?

**Efficiency and Workload**

3. What was the initial response of your team members and supervisors in implementing MATCH-AI?
4. Do you think AI has made conducting camps easier or difficult? What is your observation so far?

**Barriers and facilitators**

Van teams had option of conducting camps without MATCH-AI as well

4. What made them decide that a camp should be done using MATCH-AI?
5. What made them decide not use locations of MATCH-AI?
6. Did you get complains on difficulties in locating different camp sites selected by MATCH-AI? If so, what kind of difficulties?

**Improving Camp Yields and Performance**

Since you have been implementing MATCH AI for a while now.

4. In your opinion, what activities or strategies can help improve the yield and performance of the camps conducted through AI spot identification? What factors make a camp more successful in finding cases?
5. After MATCH AI: What is the turnover of people in camps on average? How many cases per camp do you get on average?
6. Notable meetings: who does it, how is it done? when, how, why… the turnover?

**Digital Data Usefulness**

Recently, field teams have been using digital data management tools like eCBS, while previously all data was managed on paper.

5. Which medium do you find better to use: paper-based or digital? Why or why not?
6. Is capturing digital data easy and/or useful?
7. What challenges do they face with the usage of digital tools?
8. Did you get any training or refreshers on using digital tools? How was it? Feedback.

**Staff Training**

2. What kind of additional support and/or training do they need?

**Future Use and Adaptation**

2. Would you like to incorporate long-term adoption of MATCH AI? Why or why not?

**Further Information**

2. Is there anything/something you would like to share that we haven’t asked?

**Additional Notes:**

- **Context and Background Information:**

The primary focus of this session is to understand the challenges healthcare workers face when using the MATCH AI technique for ACF camp site selection in Pakistan. We also aim to compare this AI-based approach with the conventional method. Understanding your experience will provide valuable insights into the practical application of both methods and help identify areas for improvement.

- **What MATCH-AI study adds:**

SPOT-TB is a pragmatic stepped wedge cluster randomized trial that will evaluate whether a targeted approach towards ACF, supported by artificial-intelligence software (MATCH-AI), can increase yields of TB cases detected. The study is being conducted on a national scale in Pakistan and is embedded within the routine operations of the implementing partners of the National TB Program. The study will provide empirical evidence for the potential benefit of targeting ACF interventions in a real-world setting, within a high-burden TB country.

- **Specific Instructions:**

Participants are encouraged to share both positive and negative experiences with the MATCH AI system. We are interested in how the system impacts your daily operations, your workflows, and overall staff efficiency. Please be candid and share as much detail as possible, as your input will guide future improvements.

- **Managing Responses and Encouraging Discussion:**

It is important to ensure that all participants feel comfortable expressing their opinions. If a participant is hesitant, gently prompt them with follow-up questions to draw out deeper insights. For example, ask about specific experiences or challenges. Ensure everyone has an opportunity to speak by keeping track of responses and directing questions to quieter participants.

- **Managing Time:**

Aim to stick to the suggested timings for each section to ensure the session remains focused. If a discussion is going off-topic or too detailed, kindly steer it back to the specific questions or themes.

**Closing Remarks:**

Thank you so much for your time and valuable contributions today. Your feedback is extremely helpful in improving the MATCH AI system and enhancing the ACF camp planning process. We will analyze the information gathered from all participants and use it to identify key areas for improvement in both the AI technique and the traditional camp site selection approach. We’ll follow up with a summary of the findings, and any significant changes or next steps will be communicated to you in due course. If you have any further thoughts after today’s session, feel free to reach out.

**Timing/Duration:**

Estimated Total Duration: 45- 60 minutes

**Participant Consent Form**

**Study Title:** Exploring Challenges and Barriers in Existing Practices for Camp Planning with a focus on site-selection of ACF camps using MATCH AI technique in Pakistan

**Research Team:**
Alina Shahid, Amna Mahfooz, Alizeh Faran, Abdullah Latif, Wasim Ahmed, Syed Mohammad Asad Zaidi, Nainan Nawaz, Tahira Ezra Reza, Faran Emmanuel

**Purpose of the Study**

This study aims to explore the challenges and barriers faced in planning and implementing ACF camps, focusing on the site selection process using the MATCH AI technique. Your participation will help us understand the perspectives of implementers, compare site selection approaches, and identify areas for improvement.

**What Your Participation Involves**

If you agree to participate, you will:

1. Take part in a focus group discussion or in-depth interview, lasting approximately 45 minutes to an hour.
2. Be asked about your role in the organization, experiences with MATCH AI, challenges faced, and your perspectives on camp planning processes. Questions regarding other matters that do not serve the purpose of this research, will not be asked.

Participation is entirely voluntary; there is no obligation to participate. If you choose not to participate, it is completely fine with the researcher. If you decide to participate, you can choose not to answer specific questions or withdraw at any time without penalty. Your decision to not participate, or to stop participating or to refuse to answer a specific question will not influence your relationship with the researcher, either now or in the future.

**Confidentiality**

1. Your responses will be anonymized and will not be linked to your identity.
2. Data collected during this study will be securely stored and used only for research purposes. The responses will be both audio recorded and handwritten.
3. Results may be published, but your identity will not be disclosed.

**Potential Risks and Benefits**

- **Risks:** There are no anticipated risks to participating in this study.
- **Benefits:** Your insights will contribute to improving ACF camp planning and implementation, potentially enhancing public health outcomes.

**Contact Information**

If you have any questions or concerns about the study, please contact:
Alina Shahid []

**Consent Statement**

I have read and understood the information provided above. I voluntarily agree to participate in this study. I understand that I may withdraw at any time without any negative consequences.

**Participant's Name**:

**Participant's Signature**:

**Date**:

**Researcher’s Name**:

**Researcher’s Signature**:

**Date**:

**Additional Consent (where applicable)**

You must seek additional consent by including check boxes or requesting additional signatures for the following:

1. **Audio Recording**

I consent to the audio recording of my interviews. Y / N

2. **Consent to use of quotes**

 I consent to the use of quotations in any final report/publications of this research. Y / N
