## Supplementary material for "Geographical targeting of active case finding for tuberculosis in Pakistan using artificial intelligence software: a qualitative study embedded within the SPOT TB trial": Consent Form

**Participant Consent Form**
**Study Title:**
Exploring Challenges and Barriers in Existing Practices for Camp Planning with a focus on site-selection of ACF camps using MATCH AI technique in Pakistan
**Research Team:**
Alina Shahid, Alizeh Faran

---

**Purpose of the Study**
This study aims to explore the challenges and barriers faced in planning and implementing ACF camps, focusing on the site selection process using the MATCH AI technique. Your participation will help us understand the perspectives of implementers, compare site selection approaches, and identify areas for improvement.

**سٹڈی کا مقصد**

اس سٹڈی کا مقصد میچ اے آئی ٹیکنیک کے ذریعے سائٹ سلیکشن کے طریقوں پر توجہ مرکوز کرتے ہوئے، اے سی ایف کیمپوں کی منصوبہ بندی اور عمل درآمد میں درپیش چیلنجز اور رکاوٹوں کا جائزہ لینا ہے۔ آپ کی شرکت ہمیں پراجیکٹ چلانے والے لوگوں کے نقطہ نظر کو سمجھنے، سائٹ سیلیکشن کے طریقوں کا موازنہ کرنے اور ممکنہ بہتری لانے کے لیے نشاندہی کرنے میں مدد دے گی۔

---

**What Your Participation Involves**
If you agree to participate, you will:
1. Take part in a focus group discussion or in-depth interview, lasting approximately 45 minutes to an hour.
2. Be asked about your role in the organization, experiences with MATCH AI, challenges faced, and your perspectives on camp planning processes. Questions regarding other matters that do not serve the purpose of this research, will not be asked.

**اس سٹڈی میں آپ کی شرکت میں کیا شامل ہے:**

اگر آپ شرکت کرنے پر رضامند ہیں، تو

1- آپ سے ایک گروپ میں بات چیت (فوکس گروپ ڈسکشن) یا تفصیل سے انٹرویو میں حصہ لینے کی درخواست ہے، جو تقریباً 45 منٹ سے ایک گھنٹہ تک جاری رہے گا۔

2- آپ کے ادارے میں آپ کے کردار، میچ اے آئی کے ساتھ تجربات، درپیش چیلنجز اور کیمپ کی منصوبہ بندی کے عمل کے بارے میں آپ کے نقطہ نظر کے بارے میں سوالات پوچھے جائیں گے۔ اس تحقیق کے مقصد سے ہٹ کر دیگر معاملات پر کوئی سوالات نہیں پوچھے جائیں گے۔

Participation is entirely voluntary; there is no obligation to participate. If you choose not to participate, it is completely fine with the researcher. If you decide to participate, you can choose not to answer specific questions or withdraw at any time without penalty. Your decision to not participate, or to stop participating or to refuse to answer a specific question will not influence your relationship with the researcher, either now or in the future.

اس میں آپ کی شرکت مکمل طور پر رضاکارانہ ہے، شرکت کرنے کی کوئی پابندی نہیں ہے۔ اگر آپ شرکت نہیں کرنا چاہتے / چاہتی، تو ریسرچر کے لیے یہ مکمل طور پر قابل قبول ہے۔ اگر آپ شرکت کا فیصلہ کرتے / کرتی ہیں، تو آپ کسی مخصوص سوال کا جواب دینے سے انکار کر سکتے / سکتی ہیں یا کسی بھی وقت بغیر کسی پابندی کے سٹڈی سے دستبردار ہو سکتے / سکتی ہیں۔ آپ کا شرکت نہ کرنے، یا شرکت روک دینے، یا کسی سوال کا جواب دینے سے انکار کرنے کا فیصلہ ریسرچر کے ساتھ آپ کے تعلقات پر کوئی اثر نہیں ڈالے گا، نہ اس وقت اور نہ ہی مستقبل میں۔

---

**Confidentiality**

1. Your responses will be anonymized and will not be linked to your identity.
2. Data collected during this study will be securely stored and used only for research purposes. The responses will be both audio recorded and handwritten.
3. Results may be published, but your identity will not be disclosed.

**رازداری**

1- آپ کے جوابات کو مکمل طور پر رازداری میں رکھا جائے گا اور آپ کی شناخت سے منسلک نہیں کیا جائے گا۔

2- اس سٹڈی کے دوران جمع کیے گئے ڈیٹا کو مکمل طریقے سے محفوظ کیا جائے گا اور صرف تحقیقی مقاصد کے لیے استعمال کیا جائے گا۔ آپ کے جوابات کی آڈیو ریکارڈنگ کی جائے گی اور ہاتھ سے لکھ کر بھی محفوظ کیا جائے گا۔

3- اس سٹڈی کے نتائج شائع کیے جاسکتے ہیں، لیکن اس میں آپ کی شناخت ظاہر نہیں کی جائے گی۔

---

**Potential Risks and Benefits**

- **Risks:** There are no anticipated risks to participating in this study.
- **Benefits:** Your insights will contribute to improving ACF camp planning and implementation, potentially enhancing public health outcomes.

**ممکنہ خطرات اور فوائد**

1- خطرات: اس سٹڈی میں شرکت کے کوئی متوقع خطرات نہیں ہیں۔

2- فوائد: آپ کے تفصیل سے دیے گئے جوابات اے سی ایف کیمپوں کی منصوبہ بندی کرنے اور ان پر عمل درآمد کو بہتر بنانے میں معاون ثابت ہوں گے، جس سے عوامی صحت کے نتائج میں بہتری آسکتی ہے۔

---

**Contact Information**

رابطہ کی معلومات

اگر آپ کو اس سٹڈی سے متعلق کوئی سوالات یا خدشات ہوں، تو آپ سی جی پی ایچ پاکستان میں ریسرچ آفیسر مس علینہ شاہد سے 051-8357603 پر یا ای میل کے ذریعے رابطہ کر سکتے / سکتی ہیں۔

---

**Consent Statement**

I have read and understood the information provided above. I voluntarily agree to participate in this study. I understand that I may withdraw at any time without any negative consequences.

**Participant's Name:** ____________________________________________

**Participant's Signature:** __________________________________________

**Date:** _____________________________

**Researcher’s Name:** ___________________________________________

**Researcher’s Signature:** ________________________________________

**Date:** _____________________________

رضامندی کا بیان

میں نے اوپر دی گئی معلومات کو پڑھ لیا ہے اور سمجھ لیا ہے۔ میں رضامندی سے اس سٹڈی میں شرکت کرنے پر راضی ہوں۔ میں سمجھتا / سمجھتی ہوں کہ میں کسی بھی وقت بغیر کسی منفی نتائج کے اس سٹڈی دستبردار ہو سکتا / سکتی ہوں۔

شرکت کنندہ کا نام________________________ شرکت کنندہ کے دستخط________________________

تاریخ________________________

ریسرچر کا نام________________________ ریسرچر کے دستخط________________________

تاریخ________________________

**Additional Consent (where applicable)**

You must seek additional consent by including check boxes or requesting additional signatures for the following:

1. Audio Recording

I consent to the audio recording of my interviews. Y / N

2. Consent to use of quotes

I consent to the use of quotations in any final report/publications of this research. Y / N

اضافی رضامندی (جہاں قابل اطلاق ہو)

آپ کو درج ذیل کے لیے اضافی رضامندی حاصل کرنا ہو گی یا اضافی دستخط طلب کرنے ہوں گے:-

1- آڈیو ریکارڈنگ

میں اپنے انٹرویو کی آڈیو ریکارڈنگ کی اجازت دیتا / دیتی ہوں۔

ہاں-----1 نہیں-----2

2- اقتباسات / جواب دہندہ کے کہے گئے الفاظ کے استعمال کی اجازت

میں اس سٹڈی کی کسی بھی فائنل رپورٹ / اشاعت میں اپنے اقتباسات (اپنے کہے گئے الفاظ) کے استعمال کی اجازت دیتا / دیتی ہوں۔

ہاں-----1 نہیں-----2
