## Supplementary material for "Geographical targeting of active case finding for tuberculosis in Pakistan using artificial intelligence software: a qualitative study embedded within the SPOT TB trial": Demographic Characteristics

**Demographic characteristics of participants**

| **ID** | **Gender** | **Province** | **Designation** | **Experience** |
| --- | --- | --- | --- | --- |
| **Focus Group Discussions** | | | | |

| F4 | Male | Federal | Program Officer ACF | 2.5 years |
| --- | --- | --- | --- | --- |
| F4 | Female | Federal | Research Officer | 3 years |
| F4 | Male | Federal | Senior Data Validation Officer | 3 years |
| F4 | Male | Federal | Coordinator ACF | 2 years |
| E8 | Male | Punjab | Project Coordinator | 3 years |
| E8 | Male | Punjab | Project Coordinator | 3 years |
| E8 | Male | Balochistan | Project Coordinator | 3 years |
| E8 | Male | Balochistan | Project Coordinator | 10 months |
| E8 | Male | Sindh | Project Coordinator | 10 months |
| E8 | Male | Federal | Project Coordinator | 3 years |
| E8 | Male | Sindh | Project Coordinator | 3 years |
| E8 | Male | KPK | Project Coordinator | 3 years |

**In-Depth Interviews**

| A1 | Male | KPK | District Field Supervisor | 3 Years |
| --- | --- | --- | --- | --- |
| A2 | Male | KPK | District Field Supervisor | 10 years |
| B1 | Male | KPK | Regional Coordinator | 13 Years |
| C1 | Male | KPK | District TB Coordinator | 6 months |
| A3 | Male | KPK | District Field Supervisor | 7 years |
| A4 | Male | KPK | District Field Supervisor | 1 year |
| B2 | Male | KPK | Regional Coordinator | 3 years |
| C2 | Male | KPK | District TB Coordinator | 8 years |
| D1 | Male | KPK | Program Manager | 8 Years |
| A5 | Male | Punjab | District Field Supervisor | 5 Years |
| A6 | Male | Punjab | District Field Supervisor | 15 Years |
| B3 | Male | Punjab | Regional Coordinator | 3 Years |
| B4 | Male | Punjab | Regional Coordinator | 3 years |
| D2 | Male | Punjab | Program Manager | 6 months |
| A7 | Male | Balochistan | District Field Supervisor | 12 years |
| C3 | Male | Balochistan | District TB Coordinator | 1.5 years |
| B5 | Female | Balochistan | Regional Coordinator | 11 months |
| A8 | Male | Balochistan | District Field Supervisor | 7 years |
| C4 | Male | Balochistan | District TB Coordinator | 15 years |
| B6 | Male | Balochistan | Regional Coordinator | 8 years |
| D3 | Male | Balochistan | Program Manager | 2.5 Years |
| D4 | Male | Sindh | Program Manager | 1.5 years |
| A9 | Male | Sindh | District Field Supervisor | 4 years |
| C5 | Male | Sindh | District TB Coordinator | 5 years |
| B7 | Male | Sindh | Regional Coordinator | 4 years |
| D5 | Male | Federal | Program Manager | 15 Years |
| A10 | Male | KPK | District Field Supervisor | 7 years |
| A11 | Male | KPK | District Field Supervisor | 10 years |
| C6 | Male | KPK | District TB Coordinator | 3 years |
| B8 | Male | KPK | Regional Coordinator | 5 years |
