## Supplementary material for "Geographical targeting of active case finding for tuberculosis in Pakistan using artificial intelligence software: a qualitative study embedded within the SPOT TB trial": STS Interpretation

| **STS Domain** | **Definition** | **Interpretation in Study** |
| --- | --- | --- |
| CULTURE | Culture refers to the shared values, norms, beliefs, assumptions, and behavioral expectations that shape how people work and interact within the organization. | In this study, Culture captures the gender norms and community-level social expectations that shaped how families engaged with TB screening camps, irrespective of how technically optimal a site was. Codes such as families not attending because of distance, restricted mobility for female patients, and the absence of communities within a reasonable radius reflect how prevailing cultural expectations around women’s mobility and household roles influenced camp attendance. This construct highlights that acceptance of, and engagement with, MATCH-AI-selected sites depended on the surrounding socio-cultural context of communities, not on locational accuracy alone. |
| PEOPLE | People represent the human element within the socio-technical system, including stakeholders, staff, patients, and community members. This construct emphasizes roles, responsibilities, interpersonal relationships, and capacity-building efforts. | In this study, People refers to the field staff, supervisors, and multi-level stakeholders (DTC, DHO, any other public officials or politicians) who implemented, oversaw, or were affected by MATCH-AI. It captures the tacit local knowledge that staff drew on during manual site selection such as gauging notables’ cooperation, ensuring family presence, and using personal contacts to trace patients which was a form of interpersonal, experience-based judgement that AI recommendations could not fully replicate. It also reflects varying levels of stakeholder engagement across the hierarchy, uneven awareness of how MATCH-AI functions and purpose of introducing it, and the top-down manner in which AI-driven decisions were communicated without adequately accounting for field-level context. |
| TECHNOLOGY | Technology encompasses the hardware, software, tools, equipment, information systems, and other technical resources used to support organizational processes and people. | In this study, Technology refers to MATCH-AI itself and the associated digital data systems used to support TB case-finding. It captures an ambivalent picture: on one hand, the AI’s capacity to draw on public registers and produce systematic, data-driven site recommendations was valued for identifying cases and locations that might otherwise have been missed; on the other, its algorithmic limitations were noted, including a tendency to repeat previously covered areas over time. This construct also reflects the added digital workload created by parallel manual and digital data entry, leading to missing or duplicate records, set against staff’s appreciation of features such as simultaneous, shared access to digital data. |
| INFRASTRUCTURE | Infrastructure includes the physical workplace, buildings, facilities, organizational structures, utilities, and other supporting resources that enable organizational work. | In this study, Infrastructure encompasses both the physical/geographic resources and the digital-technical infrastructure that enabled or constrained the practical use of MATCH-AI. Physical infrastructure challenges like long distances, difficult terrain, inaccessible or mountain-top locations, narrow streets unsuited to vans, scattered populations, and security concerns meant that a data-optimal site could still be practically unreachable. Digital infrastructure issues, including internet/sync failures, application crashes, data loss, and inconsistencies between tablet and web versions, further constrained smooth day-to-day use of the tool, underscoring how underlying infrastructure shaped the real-world feasibility of AI-generated recommendations. |
| PROCESSES | Processes are the formal and informal workflows, procedures, policies, and methods through which work is organized and performed to achieve system objectives. | In this study, Processes capture the formal and informal workflows through which TB camp site selection and case-finding were carried out, spanning both AI-assisted and manual, human-led procedures. This includes perceived procedural gains attributed to MATCH-AI, such as reduced favoritism/bias in site selection and, in some cases, improved case yield, set against process-level limitations such as the AI’s inability to guarantee a contextually appropriate or optimal location. It also encompasses established preparatory steps that remained central to the workflow regardless of the selection method used including pre-camp visits and engagement with local notables to reduce stigma and spread the word on TB. So this highlights that technology adoption should be operated alongside, rather than replaced by existing procedural practices. |
| GOALS | Goals define the purpose of the system, including its objectives, desired outcomes, and performance measures that guide organizational activities and decision-making. | In this study, Goals reflect participants’ perceptions of MATCH-AI’s intended purpose and value in strengthening TB case-finding, and the broader organizational objectives associated with its adoption. This is illustrated by an overall positive reception of the tool and optimism about its future role, together with reports of increased efficiency and more structured, data-driven camp planning that reduced workload for field staff. It also captures the training and capacity-building needs identified by staff, particularly requests for periodic refresher training, as a condition for sustaining progress toward these objectives indicating that achieving the tool’s intended goals depended as much on ongoing orientation of its users as on the technology itself. |

Ref link:https://business.leeds.ac.uk/research-stc/doc/socio-technical-systems-theory
